# Vertebral and Carotid Artery Stenosis after Irradiation Both Raise Ischemic Stroke Risk in Head and Neck Cancer Patients

**DOI:** 10.1101/2024.05.07.24307026

**Authors:** Jian-Lin Jiang, Joseph Tung-Chieh Chang, Bing-Shen Huang, Ting-Yu Chang, Pi-Shan Sung, Yi-Chia Wei, Chien-Yu Lin, Chih-Hua Yeh, Kang-Hsing Fan, Chi-Hung Liu

## Abstract

**Purpose:** To study the risk of ischemic stroke (IS) following carotid (CAS) or vertebral artery stenosis (VAS) in head and neck cancer (HNC) patients after radiation therapy (RT).

**Methods:** We included HNC patients who received RT between 2010 and 2023. They underwent regular head and neck imaging monitoring to evaluate cancer recurrence at the department of radiation-oncology and vascular examinations at the department of neurology. Patients were initially divided into nasopharyngeal carcinoma (NPC) and non-NPC groups. The primary outcome was the occurrence of IS after RT, and the secondary outcomes included the development of >50% CAS or >50% VAS after RT. Cox regression and Kaplan–Meier analyses were conducted to compare the outcomes of the study groups. Further analysis was conducted based on the presence or absence of >50% CAS or >50% VAS during the follow-up period.

**Results:** Of the 1,423 HNC patients, 19%, 6.8%, and 2.3% developed >50% CAS, >50% VAS, and IS during a 58-month follow-up. Compared with the NPC group, the non-NPC group exhibited a higher incidence of >50% CAS (25.4% vs. 10.7%, p<0.001) and >50% VAS (8.8% vs. 4.3%, p<0.001), but similar risks of IS. In patients with HNC, >50% CAS (adjusted hazard ratio [HR]=3.21, 95% confidence interval [CI]=1.53-6.71), and >50% VAS (adjusted HR=2.89, 95% CI=1.28-6.53) were both the independent predictors of IS. In the patients with NPC, >50% CAS was an independent predictor of anterior circulation IS (adjusted HR=4.39, 95% CI=1.17-16.47). By contrast, >50% VAS emerged as a predictor of posterior circulation IS in both the NPC (adjusted HR=15.02, 95% CI=3.76-60.06) and non-NPC groups (adjusted HR=13.59, 95% CI=2.21-83.46).

**Conclusion:** HNC patients with >50% CAS or >50% VAS after RT had an increased risk of IS within their corresponding vascular territory. CAS is a major predictor of IS in NPC patients, whereas VAS is a major predictor of IS in both NPC and non-NPC patients.

## Introduction

Head and neck cancers (HNC) is a major cause of cancer-related mortality in Asian.^1^ With continual advancements in treatment techniques, patients with HNC may experience improved long-term survival.^2^ Hence, improving the long-term quality of life of these patients is essential.^3^ Radiation therapy (RT) is a primary treatment for HNC, but it may be associated with post-radiation complications.^4, 5^ Ischemic stroke (IS) is a common post-RT complication that occurs at a rate of 17.32 per 1,000 person-years, with an incidence rate ranging from 10% to 34% among HNC survivors after RT.^6–10^ Studies have reported that compared with the general population, patients with HNC had a standardized stroke incidence ratio of 1.37^7^ and had a relative risk of IS ranging from 2.09 over a 5-year follow-up period to 10.1 over a 7-year follow-up period after RT.^8, 9^. Because IS may cause disability and reduce quality of life,^11, 12^ stroke prevention is essential for patients with HNC.

Carotid artery stenosis (CAS) is a well-documented regional complication that occurs after RT in patients with HNC, and it is regarded as a major cause of IS in this population.^13^ The overall prevalence rates of ≥50% CAS, ≥70% CAS, and carotid occlusion after RT have been revealed to be 25%, 12%, and 4%, respectively.^14–19^ The incidence rates of ≥50% CAS over cumulative periods of 12, 24, and 36 months after RT were also reported to be 4%, 12%, and 21%, respectively.^14^ Therefore, carotid duplex ultrasound (CDU) has been implemented for CAS surveillance in patients with HNC after RT.^20^ Compared with CAS, vertebral artery stenosis (VAS) after RT has received less clinical attention.^21^ According to a study utilizing magnetic resonance angiography (MRA), the incidence of VAS could reach 34.7% in patients with HNC after RT.^22^ With advancements in RT, the radiation doses delivered to the vertebral artery can be markedly reduced.^23^ Nevertheless, further research is warranted to determine the incidence of VAS after RT in patients with HNC. Compared with the general population, patients with CAS or symptomatic VAS may be at a higher risk of IS.^24, 25^ Due to the lack of studies with large cohorts, the relative risk of IS in patients with HNC who experience CAS or VAS after RT remains unclear. These evidence gaps are critical to the vascular surveillance and preventive strategy in HNC patients who receive RT. Accordingly, the present study used cohort data to explore the etiology of IS and compare the risk of IS occurrence in HNC patients with CAS or VAS across different cancer types.

## Materials and methods

### Patient recruitment

In 2022, a cohort registry was established within the four branches of our institution to examine the vasculopathy and stroke patterns of patients with HNC after RT (ClinicalTrials.gov identifier: NCT06111430). Patients were referred from our radio-oncology department through a non-probability convenient sampling. The present study used this registry to prospectively follow up these patients and retrospectively review their data. The study protocol was approved by the ethics and institutional review board of our hospital (202101981B0, 202200464B0, and 202400107B0). Written informed consent was obtained from all eligible patients. Using this registry, we initially screened patients with HNC who had received RT at our institution between January 1, 2010, and December 31, 2023. We excluded patients who were lost to follow-up, received both proton beam therapy (PBT) and volumetric modulated arc therapy (VMAT), or had significant cervical cranial vessel stenosis (VS)—including CAS or VAS—before RT (Figure 1). This study was conducted in accordance with the Strengthening the Reporting of Observational Studies in Epidemiology reporting guidelines.

**Figure 1:**
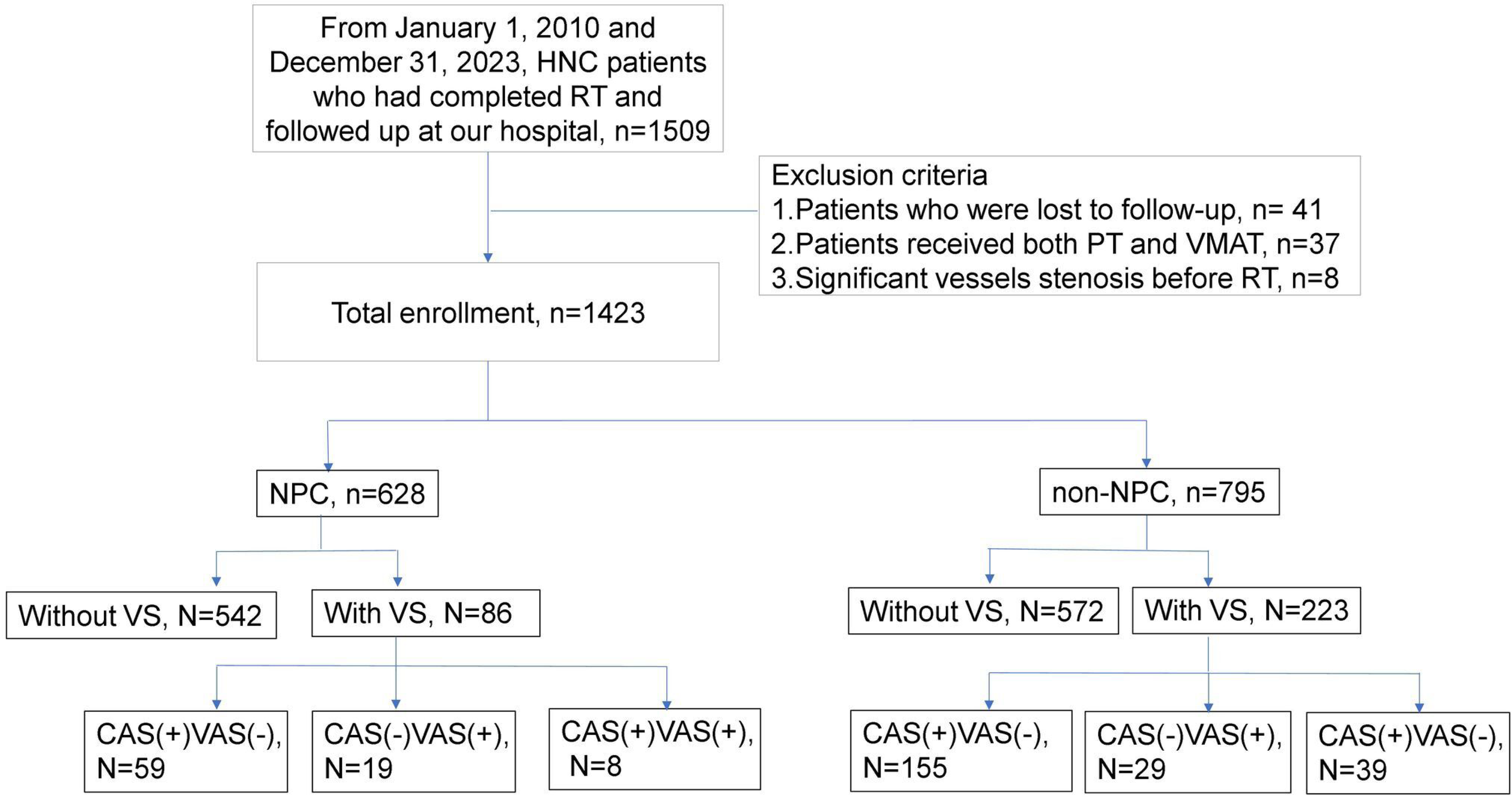
Flowchart of participant enrollment. **Abbreviations**: CAS, carotid artery stenosis; HNC, head and neck cancer; NPC, nasopharyngeal carcinoma; RT, radiation therapy; VAS, vertebral artery stenosis; VS, vessel stenosis.

Demographic data and common stroke risk factors, including hypertension, diabetes mellitus, dyslipidemia, atrial fibrillation, and cigarette smoking, were recorded. Data on medication use, including antithrombotic agents and statins, were collected. Cancer types, tumor–node–metastasis stages, and treatment methods for HNC, including RT, surgical treatment, and chemotherapy, were documented. Details on RT, including the applied RT method (PBT or VMAT), total cumulative dose of RT, and the time interval between the last RT fraction and latest clinical follow-up, were recorded.

### RT methods

Radiation treatment plans for PBT and VMAT were established using an Eclipse planning system (version 13.7; Varian Medical Systems, Palo Alto, CA, USA). These plans adhered to the same dose constraints and optimization algorithms. A relative biological equivalent value of 1.1 was assumed for PBT. The prescribed doses for postoperative RT and primary RT were 6,000–6,600 cGy delivered in 30–33 fractions and 6,996 cGy delivered in 33 fractions, respectively, over a period of 6 to 7 weeks (five fractions per week). Any deviation in total treatment time exceeding 5 days beyond the scheduled duration was considered a major violation.^26^

### Follow-up strategy

Patients underwent regular follow-up assessments at the department of radiation oncology every 6 months and were referred to the department of neurology for neurovascular complication monitoring after RT. At the radiation oncology visits, head and neck computed tomography (CT) and magnetic resonance imaging (MRI) scans were performed 3 months after RT and then every 6–12 months to monitor potential cancer recurrence. A multidetector CT scanner was used to produce thin-slice images (3-mm thickness) through multiplanar reformation. Head and neck MRI was performed using 1.5- or 3.0-T scanners equipped with a standard head and neck coil. These scans were obtained in the axial plane with a section thickness of 5 mm and intersection gap of 2.5 mm and in the sagittal and coronal planes with a section thickness of 4 mm and gap of 1 mm. At the neurology visits, CDU studies were scheduled every 1 to 2 years to monitor the development of CAS or VAS. For patients who had >50% CAS or VAS on follow-up CT or MRI axial images^27–29^ and patients who had >50% CAS or VAS based on B-mode findings^30^ or hemodynamic criteria^31–33^ in serial CDU studies, additional diagnostic procedures such as CT angiography or MRA were conducted to confirm the severity of vascular stenosis.

### Outcomes

The primary outcome was the occurrence of IS after RT. An IS was defined as a symptomatic infarction with corresponding findings on brain CT or MRI scans. The secondary outcomes were the development of >50% CAS, the development of >50% VAS, and mortality after RT. Vascular outcomes were validated by an experienced neuroradiologist (CHY) and two vascular neurologists (CHL and JLJ).

### Grouping

The patients were divided into two groups according to the type of cancer: a nasopharyngeal carcinoma (NPC) group and a non-NPC group (Figure 1). The non-NPC group comprised patients with cancer of the oral cavity, oropharynx, larynx, or hypopharynx. To determine the correlation between vascular stenosis and the risk of IS, the patients were further grouped depending on the presence or absence of significant VS after RT. Significant VS was defined as the occurrence of either >50% CAS or >50% VAS during follow-up.

### Statistical analysis

All statistical analyses were conducted using R software (version 4.3.1; Beagle Scouts). Kolmogorov–Smirnov tests were conducted to evaluate continuous variables, and chi-square tests were used to evaluate categorical variables, such as dyslipidemia, hypertension, diabetes mellitus, alcohol consumption, and cigarette smoking. Continuous variables, including baseline glycated hemoglobin, low-density lipoprotein cholesterol, and serum creatinine levels, were evaluated using either a two-sample t test (age) or the Mann–Whitney U test (radiation dose and follow-up duration). Cox regression and Kaplan–Meier analyses were conducted to compare the risk of IS between the NPC and non-NPC groups, and multivariate analyses were conducted to adjust for covariates such as CAS, VAS, hypertension, diabetes mellitus, and dyslipidemia. Cox regression and Kaplan–Meier analyses were also used to compare the risk of IS in the NPC and non-NPC groups with and without significant VS. Survival curves were adjusted for hypertension. A p value of <0.05 was considered statistically significant.

## Result

### Patient recruitment and baseline characteristics

We initially included 1,509 patients from the cohort registry. Among these patients, 41 had missing follow-up data, 37 had received both PBT and VMAT, and 8 had >50% CAS or >50% VAS before RT and were thus excluded. Accordingly, we finally contained 1,423 HNC patients for analysis. Of them, 628 (44%) patients were classified into the NPC, and 795 (56%) were classified into the non-NPC group. The median follow-up duration was 58 months. In the NPC group, 86 (13.7%) patients had significant VS, of whom 59 (68.6%) had >50% CAS only, 19 (22.1%) had >50% VAS only, and 8 (9.3%) had both >50% CAS and >50% VAS. In the non-NPC group, 223 (28.1%) patients had significant VS, of whom 155 (69.5%) had >50% CAS only, 29 (13%) had >50% VAS only, and 39 (17.5%) had both >50% CAS and >50% VAS (Figure 1).

Compared to the non-NPC group, patients of the NPC group were younger, less male predominance, had lower frequency of hypertension, smoking, statin, and antiplatelet use. Additionally, the NPC group had a higher frequency of PBT treatment and higher radiation dose. (Supplementary Table 1).

In the NPC group, patients with significant VS were older (56 vs. 52 years, p=0.002), were predominantly men (86% vs. 76%, p=0.042), had a higher incidence of hypertension (24% vs. 14%, p=0.013), and had a higher frequency of statin (76% vs. 30%, p<0.001), and antiplatelet use (62% vs. 8.3%, p<0.001) compared with their counterparts. They also had a lower frequency of PBT treatment (9.3% vs. 30%, p<0.001), and a longer follow-up duration (95 vs. 57 months, p<0.001). In the non-NPC group, patients with VS were also older (62 vs. 58 years, p<0.001), were predominantly men (94% vs. 85%, p<0.001), had a higher incidence of hypertension (26% vs. 19%, p=0.047), and had a higher frequency of statin (72% vs. 33%, p<0.001), and antiplatelet use (62% vs. 10%, p<0.001) compared with those without VS. They also had a lower frequency of PBT treatment (4.8% vs. 13%, p<0.001) and a longer follow-up duration (78 months vs. 47 months, p<0.001; Table 1).

**Table 1.** Baseline characteristic of NPC and non-NPC patients with and without significant vessel stenosis.

|  | Total,<br>n=1,423 | NPC, n=628 |  |  | Non-NPC, n=795 |  |  | NPC vs. non-NPC<br>P value |
| --- | --- | --- | --- | --- | --- | --- | --- | --- |
|  |  | W/O VS, N=542 | With VS, N=86 | P value | W/O VS, N=572 | With VS, N=223 | P value |  |
| Age (years) | 57 (49, 64) | 52 (44, 59) | 56 (50, 63) | 0.002* | 58 (51, 65) | 62 (56, 67) | <0.001* | <0.001* |
| Male (%) | 1,181 (83%) | 413 (76%) | 74 (86%) | 0.042* | 484 (85%) | 217 (94%) | <0.001* | <0.001* |
| Diabetes mellitus (%) | 163 (11%) | 56 (10%) | 12 (14%) | 0.315 | 70 (12%) | 25 (11%) | 0.574 | 0.509 |
| Hypertension (%) | 266 (19%) | 76 (14%) | 21 (24%) | 0.013* | 110 (19%) | 59 (26%) | 0.047* | 0.005* |
| Dyslipidemia (%) | 204 (14%) | 69 (13%) | 12 (14%) | 0.753 | 82 (14%) | 42 (18%) | 0.172 | 0.173 |
| Atrial fibrillation | 9 (0.7%) | 2 (0.4%) | 1 (1.2%) | 0.358 | 4 (0.9%) | 2 (0.9%) | >0.999 | 0.527 |
| Smoking (%) |  |  |  | 0.058 |  |  | 0.163 | <0.001* |
| No | 803 (56%) | 386 (71%) | 52 (60%) |  | 274 (48%) | 96 (42%) |  |  |
| Yes | 188 (13%) | 40 (7.4%) | 12 (14%) |  | 90 (16%) | 47 (20%) |  |  |
| Quit | 432 (30%) | 116 (21%) | 22 (26%) |  | 208 (36%) | 88 (38%) |  |  |
| Proton therapy (%) | 256 (18%) | 164 (30%) | 8 (9.3%) | <0.001* | 74 (13%) | 11 (4.8%) | <0.001* | <0.001* |
| Radiation dose. (cGy) | 6,996(6,600, 6,996) | 6,996(6,996, 7,000) | 6,996(6,996, 7,200) | 0.569 | 6,744(6,600, 6,996) | 6,600(6,600, 6,996) | 0.104 | <0.001* |
| Statin use (%) | 577 (40%) | 162 (30%) | 65 (76%) | <0.001* | 186 (33%) | 166 (72%) | <0.001* | 0.003* |
| Antiplatelet use (%) | 558 (39%) | 45 (8.3%) | 53 (62%) | <0.001* | 58 (10%) | 144 (62%) | <0.001* | <0.001* |
| Follow-up (months) | 58 (30, 103) | 57 (29, 106) | 95 (64, 144) | <0.001* | 47 (25, 79) | 78 (48, 123) | <0.001* | 0.001* |

**Abbreviations:** cGy; centigray; NPC, nasopharyngeal carcinoma; VS, vascular stenosis; W/O, without

### Primary and secondary outcomes

As presented in Table 2, 33 (2.3%) patients experienced an IS after RT. In the NPC group, 10 patients (11.6%) with significant VS experienced an IS, whereas only 7 patients (1.3%) without VS experienced an IS. In the non-NPC group, patients with significant VS exhibited a higher incidence of IS compared with those without VS (4.8% vs. 0.9%, p<0.001).

**Table 2.**
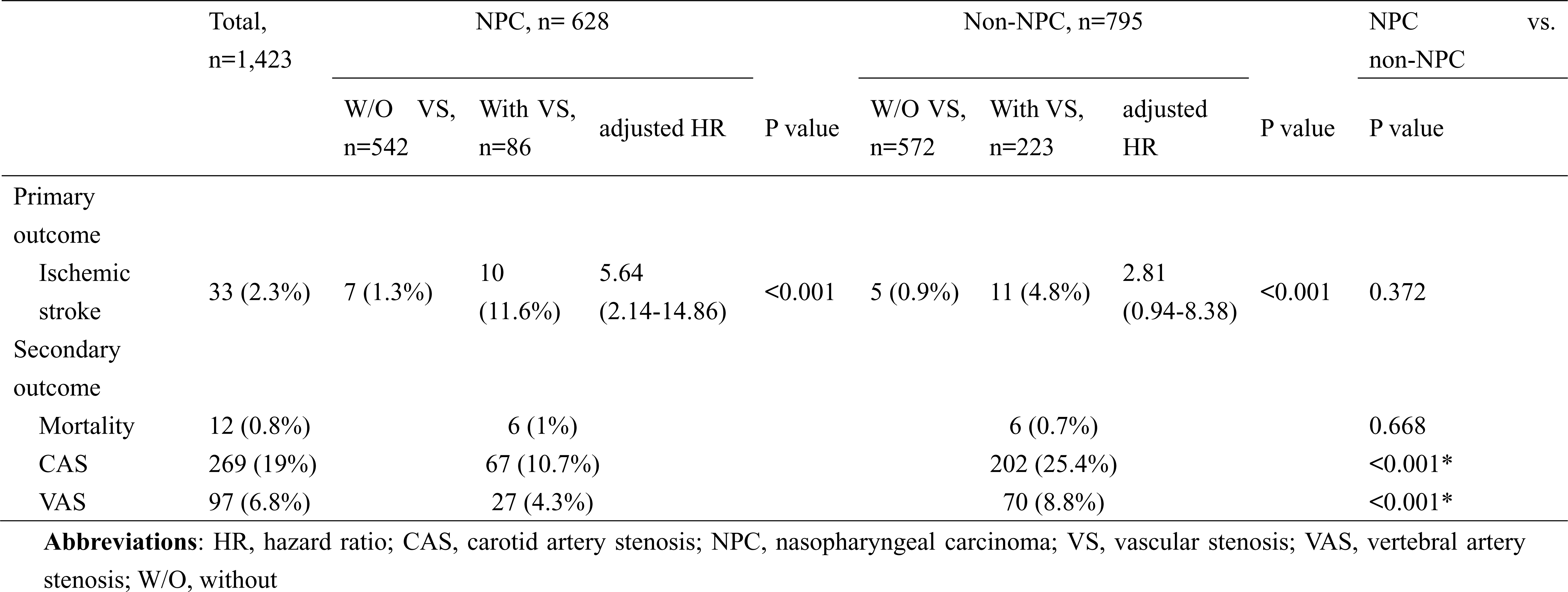
Primary and secondary outcomes of this study.

Among all HNC patients, 269 (19%) and 97 (6.8%) patients developed >50% CAS and >50% VAS, respectively. Compared with the NPC group, the non-NPC group exhibited a higher incidence of >50% CAS (25.4 % vs. 10.7 %, p<0.001) and >50% VAS (8.8 % vs. 4.3%, p<0.001). Nevertheless, no statistically significant difference was observed in the risk of IS or rate of mortality between the two groups (Table 2 and Supplementary Figure 1).

### Survival and multivariate analysis

Compared with the NPC group, the non-NPC group was more prone to develop significant VS (hazard ratio [HR]=2.94, 95% confidence interval [CI]=2.29-3.81; Supplementary Figure 2A), >50% CAS (HR =3.41, 95% CI=2.57-4.54; Supplementary Figure 2B), and >50% VAS (HR=2.45, 95% CI=1.57-3.84; Supplementary Figure 2C), as indicated by Kaplan–Meier analysis. Regarding the influence of significant VS on IS occurrence, only NPC patients with significant VS were at a high risk of IS (HR=5.93, 95% CI=2.25-15.63; Figure 2A), as indicated by Kaplan–Meier analysis. No statistically significant difference was observed in the risk of IS in non-NPC patients with VS (Figure 2B). After common vascular risk factors were adjusted for, patients with significant VS in the NPC group were at a significantly high risk (HR=5.64, 95% CI=2.14-14.86; Supplementary Figure 3A), whereas those in the non-NPC group were found to be at a similar risk (p=0.1) of IS after RT (Supplementary Figure 3B).

**Figure 2:**
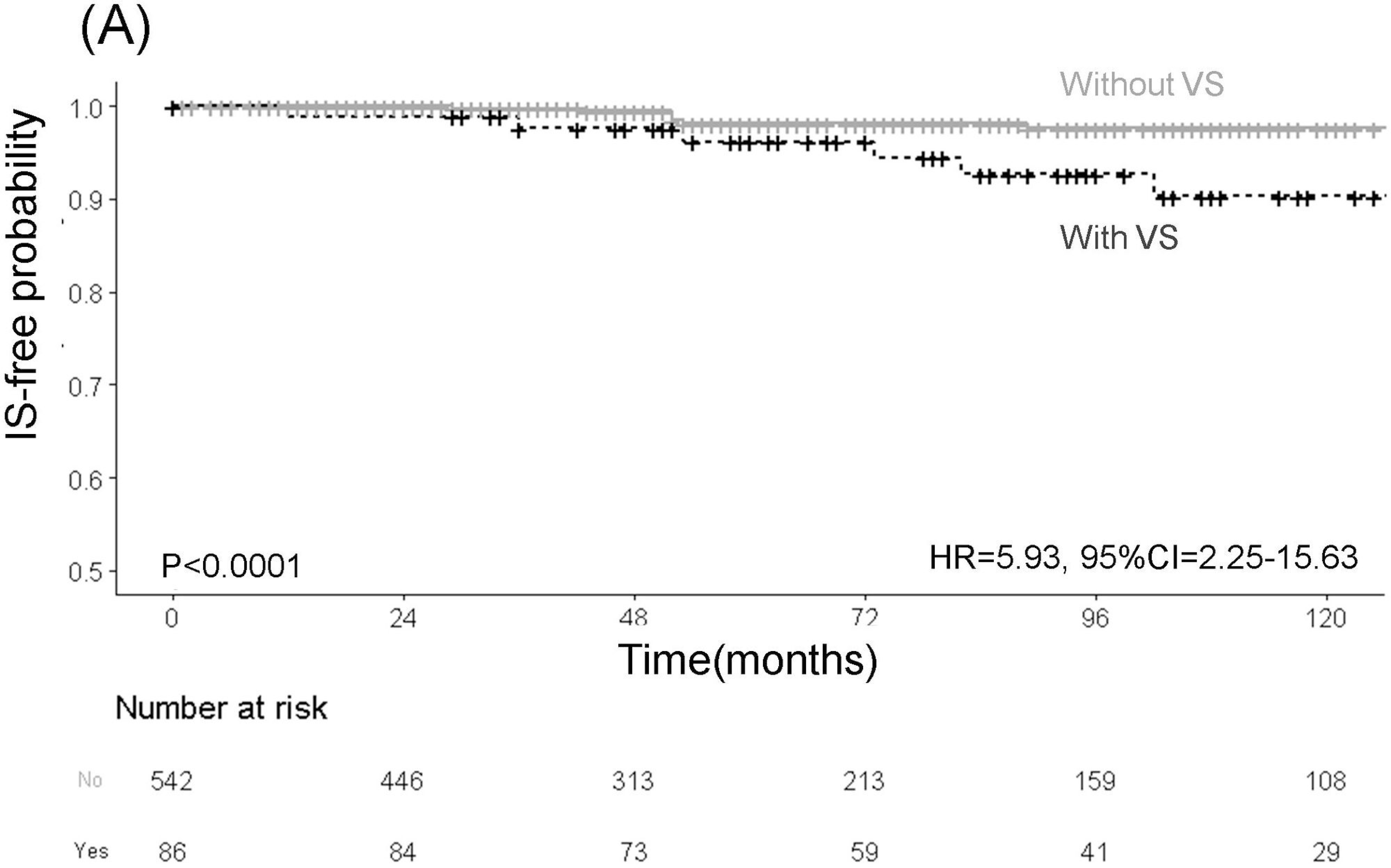
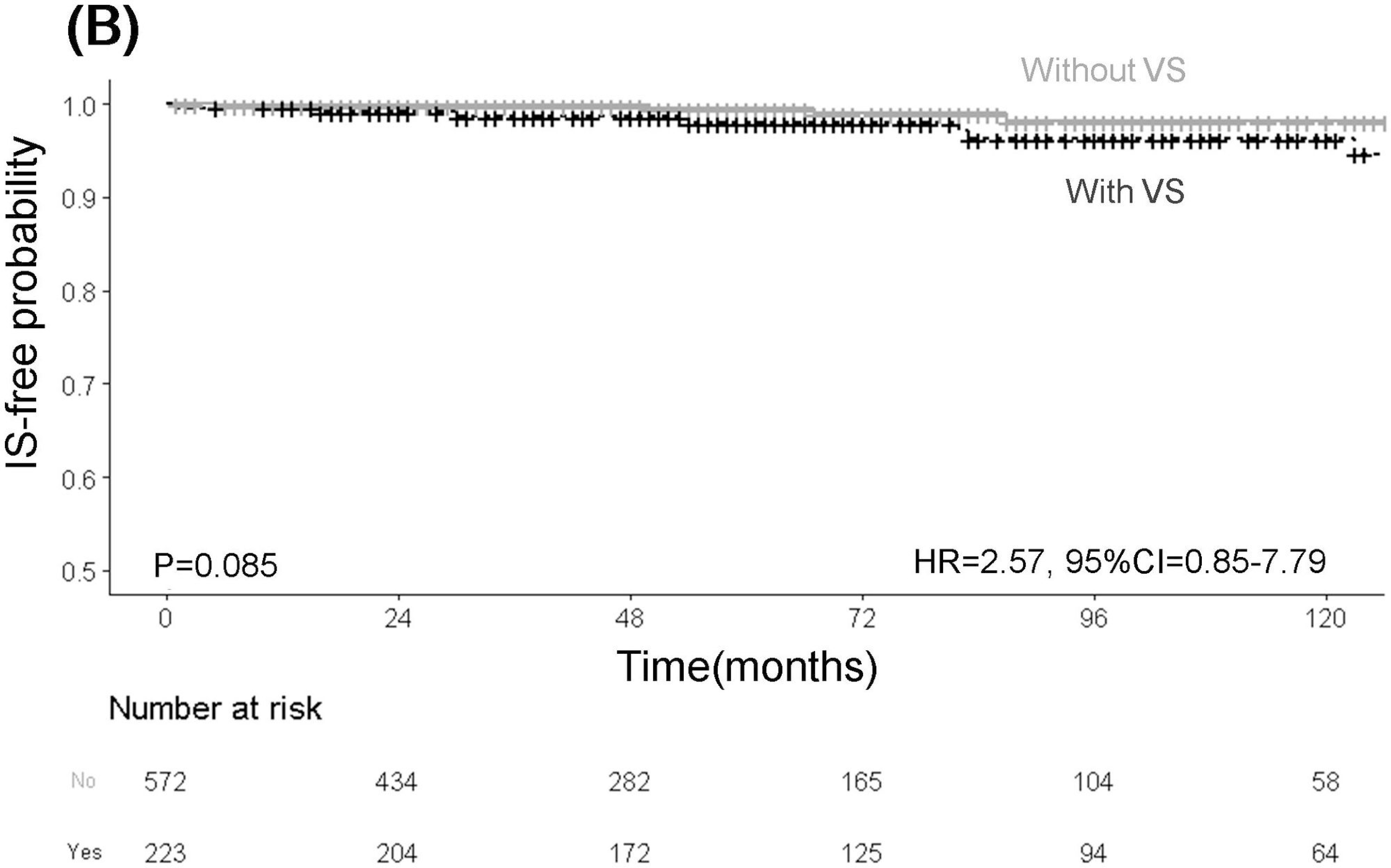
Comparison of cumulative IS-free rates in NPC (A) and non-NPC (B) patients with and without VS through Kaplan–Meier analysis. The figure depicts a high risk of IS among NPC patients with VS (A) but a comparable risk of IS among non-NPC patients with and without VS. **Abbreviations**: NPC, nasopharyngeal carcinoma; IS, ischemic stroke; VS, vessel stenosis.

Our multivariate analysis revealed that >50% CAS (adjusted HR=3.21, 95% CI=1.53-6.71), and >50% VAS (adjusted HR=2.89, 95% CI=1.28-6.53) were independent predictors of IS in patients with HNC. Specifically, >50% CAS (adjusted HR=4.22, 95% CI=1.52-11.71) was an independent predictor of IS in NPC patients, whereas >50% VAS (adjusted HR=3.4, 95% CI=1.13-10.23) was an independent predictor of IS in non-NPC patients (Table 3).

**Table 3.** Multivariate analysis results of the predictors of ischemic stroke.

|  | All HNC |  | NPC |  | Non-NPC |  |
| --- | --- | --- | --- | --- | --- | --- |
|  | adjusted HR | P value | adjusted HR | P value | adjusted HR | P value |
| Carotid artery stenosis | 3.21(1.53-6.71) | 0.002* | 4.22(1.52-11.71) | 0.006* | 2.74(0.9-8.38) | 0.08 |
| Vertebral artery stenosis | 2.89(1.28-6.53) | 0.011* | 2.95(0.84-10.36) | 0.09 | 3.4(1.13-10.23) | 0.03* |
| Hypertension | 2.01(0.87-.4.65) | 0.1 | 1.31(0.3-5.69) | 0.72 | 1.36(0.25-7.37) | 0.71 |
| Diabetes mellitus | 1.44(0.51-4.06) | 0.49 | 2.22(0.63-7.82) | 0.21 | 1.99(0.62-6.44) | 0.25 |
| Dyslipidemia | 0.48(0.15-1.48) | 0.2 | 0.72(0.14-3.64) | 0.69 | 0.41(0.08-2.11) | 0.29 |

**Abbreviations:** HNC, head and neck cancer; NPC, nasopharyngeal carcinoma

### IS patterns, and risks of IS attributing to the stenotic vessels

In accordance with the TOAST classification of IS, 60.6% of the cases were attributed to large artery atherosclerosis, and 30.3% and 6% of the cases were attributed to small vessel and cardiogenic causes, respectively. Among 18 (54.5%) and 14 (42.4%) patients with anterior and posterior circulation infarction, 60% and 57.1%, respectively, had large artery atherosclerosis (Supplementary Tables 2 and 3). NPC patients with >50% CAS exhibited a relatively high risk of IS within the corresponding vascular territory (adjusted HR=4.39, 95% CI=1.17-16.47; Figure 3A). Of notes, both NPC (adjusted HR=15.02, 95% CI=3.76-60.06; Figure 3C) and non-NPC (adjusted HR=13.59, 95% CI=2.21-83.46; Figure 3D) patients with >50% VAS were at a relatively high risk of IS in posterior circulation.

**Figure 3:**
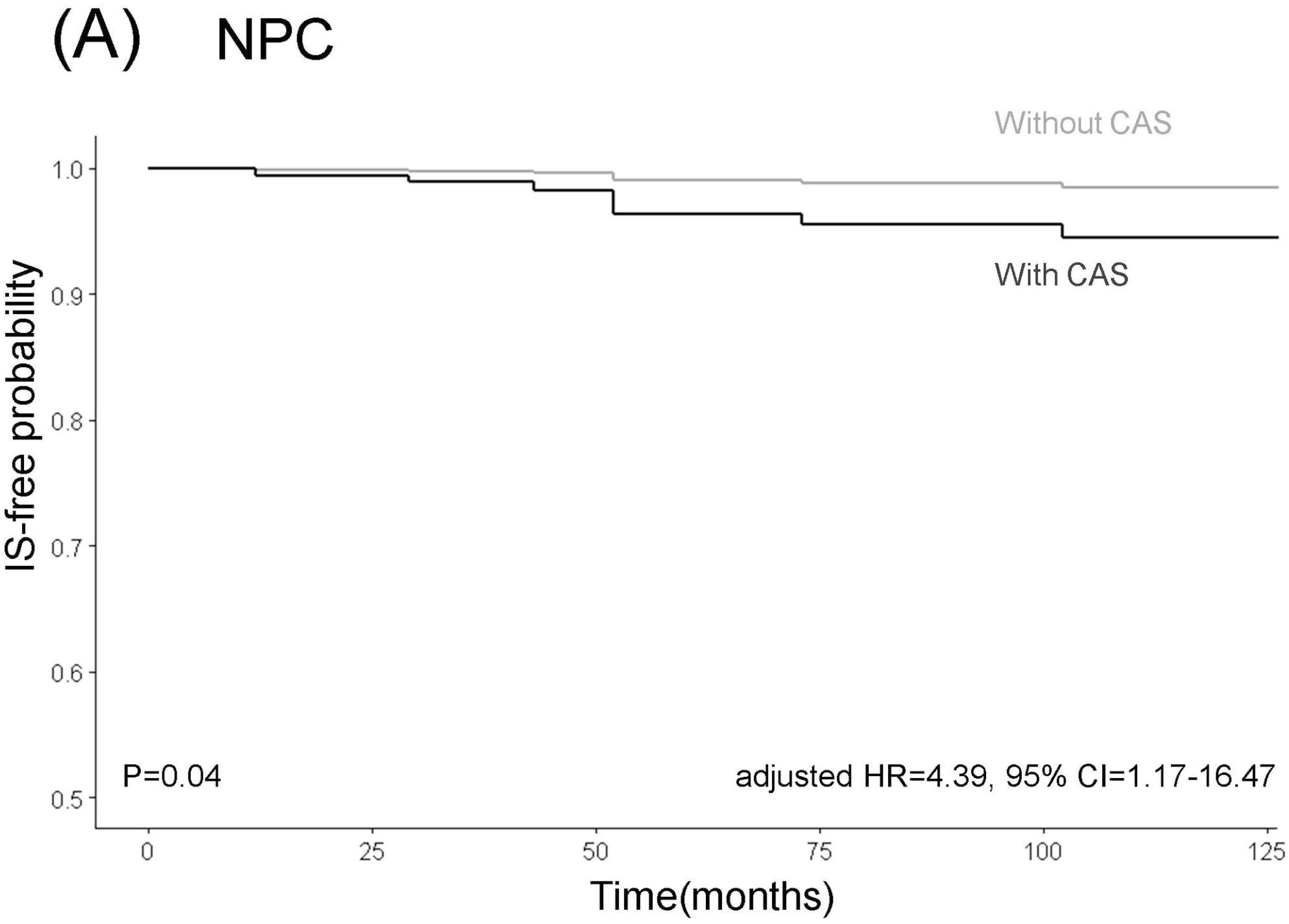
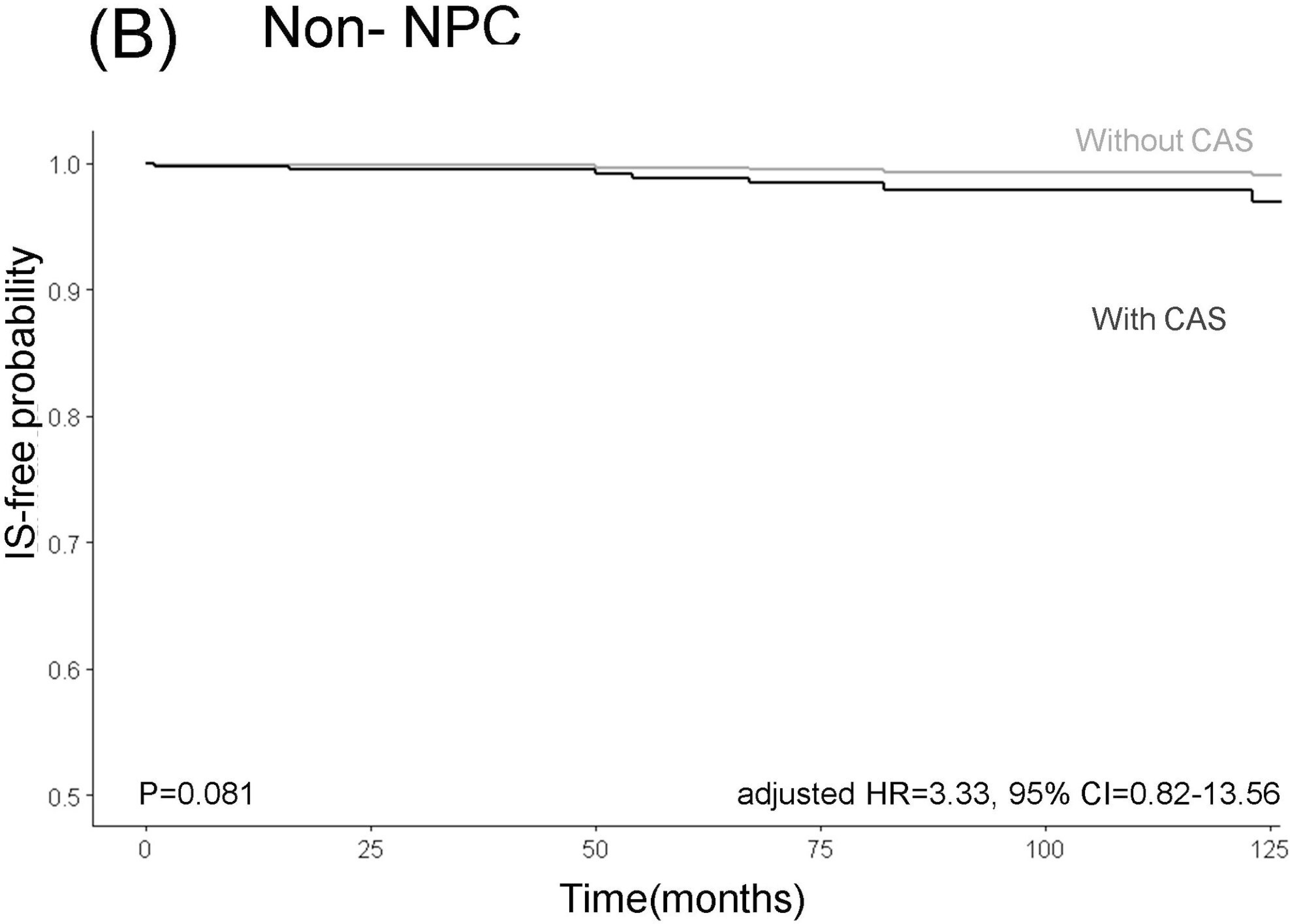
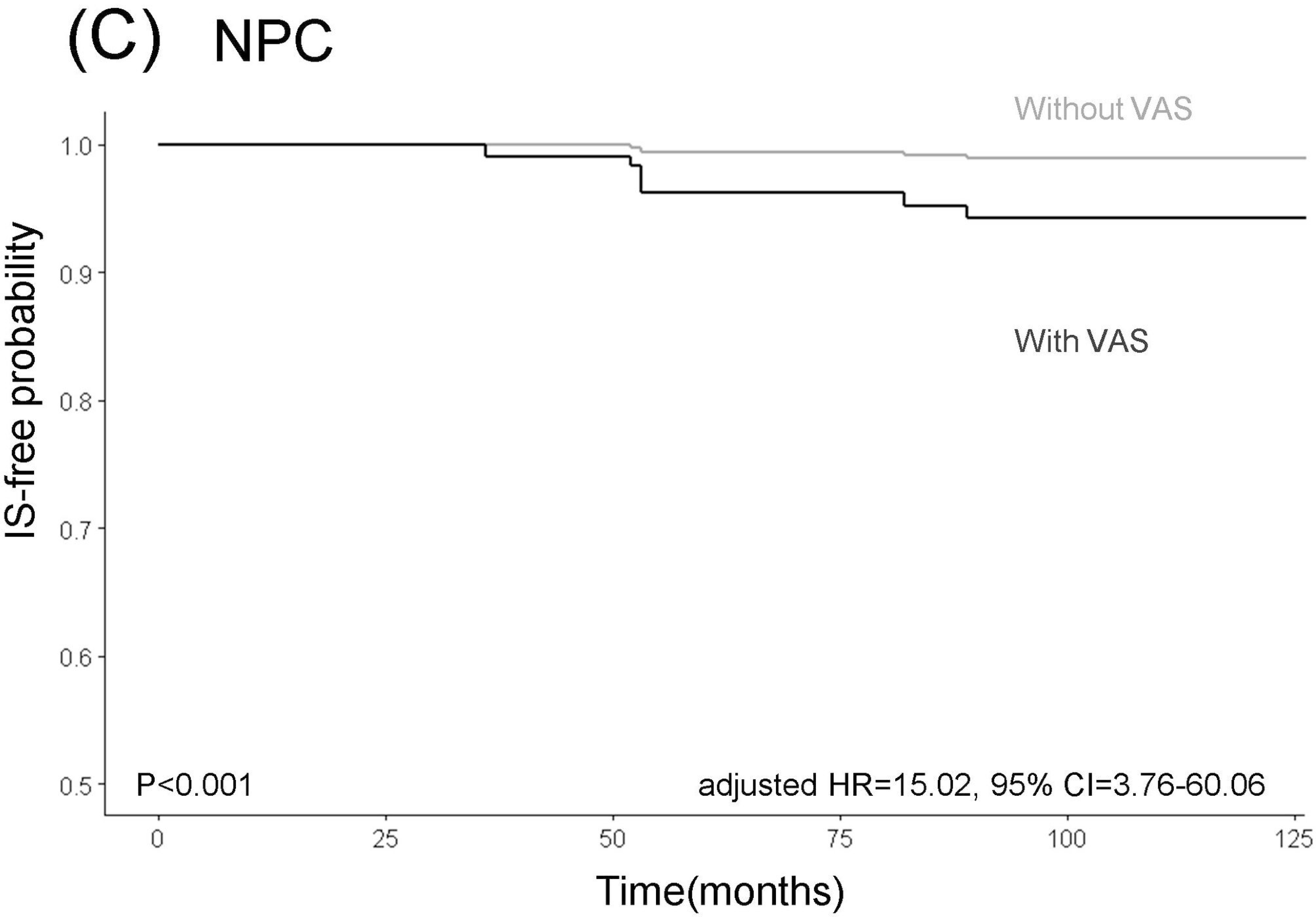
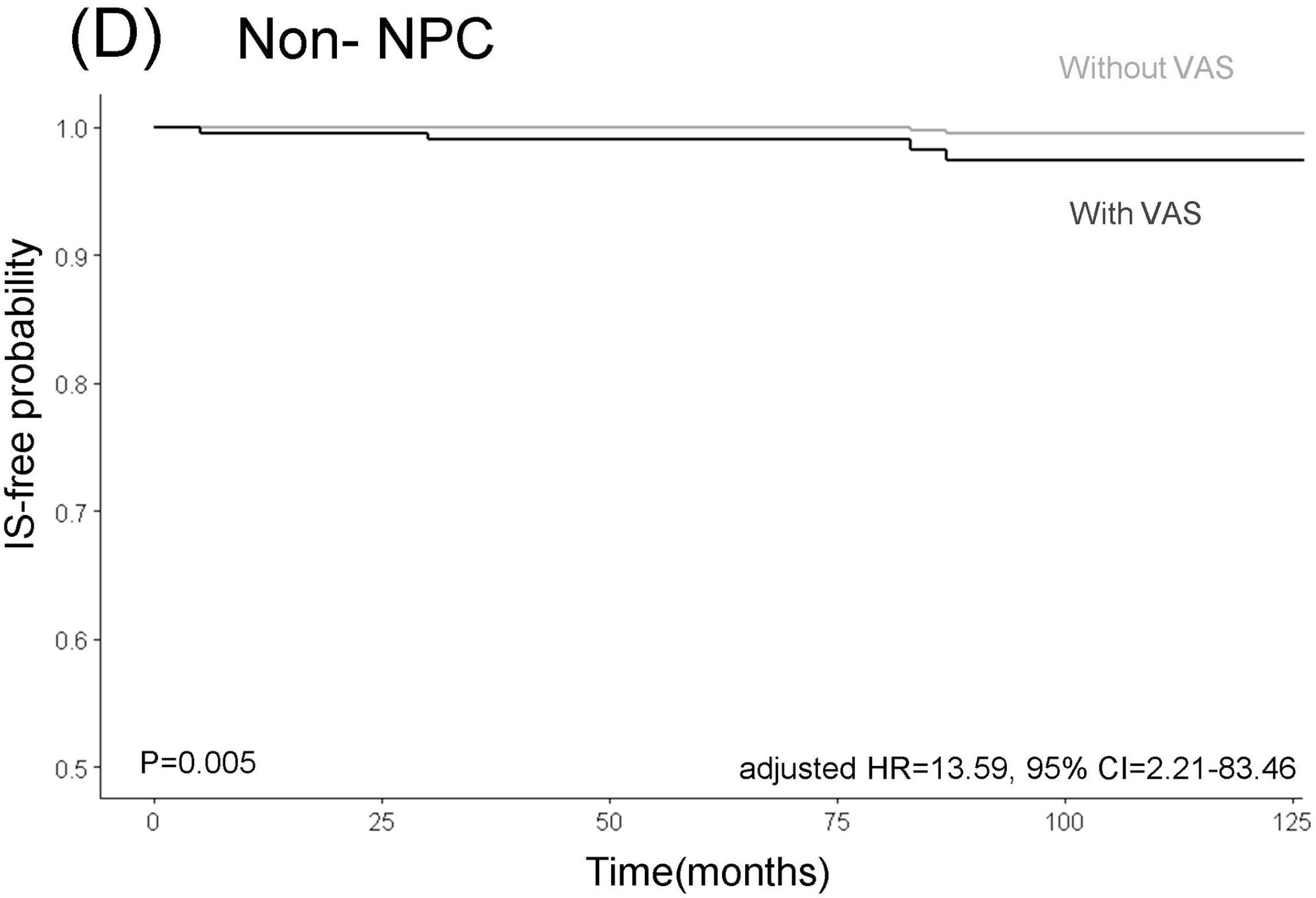
Adjusted cumulative IS-free rates in anterior circulation in NPC (A) and non-NPC (B) patients with and without CAS. Adjusted cumulative IS-free rates in posterior circulation in NPC (C) and non-NPC (D) patients with and without VAS. The figure depicts a higher risk of IS among NPC patients with >50% CAS (A) but a slightly higher IS risk among non-NPC patients (B). During follow-up, both NPC (C) and non-NPC (D) patients with >50% VAS were at an elevated risk of IS. **Abbreviations**: CAS, carotid artery stenosis; NPC, nasopharyngeal carcinoma; IS, ischemic stroke; VAS, vertebral artery stenosis.

## Discussion

Our results disclosed that among all HNC patients, 19% and 6.8% developed >50% CAS and >50% VAS during follow-up. Overall, 2.3% of our HNC patients developed IS after RT. Notably, both the >50% CAS and >50% VAS were independent risk factors for IS occurrence in the corresponding vascular territories in these patients. In addition, occurrence of >50% CAS could have higher influence on the subsequent IS in NPC patients, while occurrence of >50% VAS may have higher impact on IS in non-NPC patients. Therefore, presence of >50% VAS could be as important as >50% CAS and should be carefully monitored after RT.

Although IS commonly occurs in patients who have received RT, few studies have examined the etiology and vascular territory of IS.^6–10^ Moreover, CAS is also a common complication after RT.^14–19^ In general population, the annual IS risk was 0.4% and 0.5% for patients with 50% to 99% and 70% to 99% in asymptomatic CAS.^34, 35^ In another study, the incidence of IS associated with ipsilateral asymptomatic CAS was projected to reach 4.7% over a 5-year period.^24^ On the contrary, a study revealed that the 5-year rate of IS in patients with symptomatic CAS ranged from 19% to 21%.^36^ Therefore CAS screening has been proposed previously to mitigate the IS occurrence in HNC patients after RT. Although CAS is presumably the primary cause of IS in patients with HNC after RT, few studies have examined the long-term effects of IS in these patients after the development of CAS. Through our cohort, we observed a considerably high incidence of IS after the development of CAS in the NPC group. By contrast, the incidence of IS in non-NPC patients was close to that observed in the general population. These findings underscore the clinical importance of CAS screening in patients with NPC after RT.

We discovered similar incidence rates of IS but differences in the rates of CAS-associated IS between the NPC and non-NPC groups after RT. These findings demonstrate the importance of carefully evaluating the mechanism of IS in non-NPC patients. We also observed an increased risk of VAS-associated IS in the non-NPC group after RT. A previous study indicated that NPC patients with VAS were at a significantly increased risk of IS after RT. ^21^ Furthermore, we determined that the non-NPC group was particularly vulnerable to experiencing an IS following VAS after RT, in contrast to the NPC group. A study reported that the incidence of VAS after RT was approximately 34.7% in patients with HNC.^22^ Another study involving 3,717 patients with atherosclerotic arterial disease revealed that 7.6% of the patients had asymptomatic VAS.^37^ In the general population, approximately 20% of posterior circulation infarction cases may be attributable to VAS.^38^ In this study, we discovered that both non-NPC and NPC patients with >50% VAS were at relatively high risks of IS. Although the literature primarily emphasizes the importance of CAS surveillance after RT,^39^ our findings demonstrate the importance of VAS detection in patients with HNC.

Compared to extracranial CAS, the utility of CDU in identifying extracranial VAS may be limited.^40^ A previous study revealed the sensitivity of CDU for VAS diagnosis was only 70.2%, whereas the sensitivity levels of MRA and CT angiography for VAS diagnosis were 93.9% and 100%, respectively.^41^ Our findings suggest that regular evaluation of VAS should be incorporated into CT and MRI protocols for HNC patients during posttreatment follow-ups. Previous studies have indicated an association between statin use and a low risk of IS in HNC patients after RT.^42–44^ A study indicated that oral antithrombotic therapy did not significantly mitigate the risk of IS.^45^ In this study, we discovered that NPC patients with CAS and non-NPC patients with VAS were at elevated risks of IS. Moreover, a large proportion of patients with CAS or VAS had received statin or antiplatelet treatment. Further research is warranted to determine whether these management protocols can reduce the incidence of IS when >50% CAS or >50% VAS is detected during follow-up.

This study has several limitations. First, vascular status was evaluated using CT or MRI protocols tailored for cancer follow-up, which may have influenced the accuracy of CAS or VAS diagnosis. Although CT or MRA scans were scheduled to confirm VS in certain patients, selection bias may occur. Second, not all patients with HNC treated at our institution were included in our registry; those who were lost to follow-up may have represented a source of selection bias. Third, patients who experienced an IS at other hospitals before enrollment may have represented a source of recall bias, presumably resulting in an underestimation of the incidence of IS in this study. Fourth, this study was conducted at a single institution. Although our institution has four branches and is regarded as one of the largest centers providing PBT and treatment for HNC in Taiwan, our findings may not be generalizable to individuals from other countries. Fifth, the incidence of IS was low in our study. Therefore, future studies with larger sample sizes and longer follow-up durations are warranted to determine the influence of different RT methods, cancer types, and preventive medications on the occurrence of IS.

## Conclusion

HNC patients with >50% CAS or >50% VAS after RT both have a relatively high risk of IS in their corresponding vascular territory. CAS is a major predictor of IS in NPC patients, and VAS is a major predictor of IS in both NPC and non-NPC patients. Vascular evaluation after RT should include both the carotid and vertebral arteries because they are equally essential. Further multicenter prospective research is warranted to determine the influence of preventive interventions on the occurrence of IS when CAS or VAS is detected.

## Declarations

### Ethics Approval

The study was approved by the Ethics Institutional Review Board of Chang Gung Memorial Hospital (202101981B0, 202200464B0, and 202400107B0).

### Data Availability

The data sets generated in this study are available from the corresponding author upon reasonable request.

### Author’s contributions

All authors have read and approved the manuscript

Conception and design: CHL, JLJ, TCC

Analysis and interpretation: JLJ, CHL, KHF, BSH, CYL, YCW, PSS

Data collection: JLJ, CHL, TCC, KHF, CYL, TYC, YCW, CHY

Writing the article: JLJ, CHL, TCC

Critical revision of the article: JLJ, KHF, BSH, PSS, CHL, TYC

Final approval of the article: CHL, TCC

Statistical analysis: JLJ, CHL

Obtained funding: CHL

Overall responsibility: CHL

## Acknowledgments

The authors thank Chang Gung Memorial Hospital, the Ministry of Science and Technology of Taiwan, Ms. Karen Kang, and Ms. Elaine Shinwei Huang for their administrative efforts.

## Abbreviation

CAS: carotid artery stenosis
CI: confidence interval
CT: computed tomography
CDU: carotid duplex ultrasound
HNC: head and neck cancer
HR: hazard ratio
IS: ischemic stroke
NPC: nasopharyngeal carcinoma
MRA: magnetic resonance angiography
MRI: magnetic resonance imaging
PBT: proton beam therapy
RT: radiation therapy
VAS: vertebral artery stenosis
VMAT: volumetric modulated arc therapy
VS: vessel stenosis

